# Directionally opposing effects of a shared immune genetic signature on atopic dermatitis and glioblastoma: integrative insights from single-cell and clustered Mendelian randomization analyses

**DOI:** 10.64898/2026.05.01.26352272

**Authors:** Xiaomei Chen, Hua Ye, Jizhen Yang, Tao Qu

## Abstract

**Background:** Epidemiological studies have consistently documented an inverse association between atopic dermatitis (AD) and glioblastoma (GBM), yet the immunogenetic mechanisms underlying this paradox remain elusive. We hypothesized that distinct immune subsets driven by shared genetic variants exhibiting antagonistic pleiotropy may explain this relationship.

**Objective:** To dissect the immunogenetic basis underlying the inverse association between AD and GBM by integrating single-cell transcriptomics and clustered Mendelian randomization, and to identify shared immune subsets and genetic variants exhibiting antagonistic pleiotropy that may explain this epidemiological paradox.

**Methods:** We integrated single-cell RNA sequencing (scRNA-seq) of publicly available datasets from AD skin (GSE153760) and GBM tumors (GSE256490) with genome-wide association study (GWAS) summary statistics. Disease-specific immune cell subsets were identified, and pathway enrichment was conducted on marker genes. Clustered Mendelian randomization (MR-Clust) was applied to detect heterogeneous causal effects, followed by drug target enrichment analysis using the DGIdb database.

**Results:** scRNA-seq revealed that Th2A cells were the predominant pathogenic subset in AD lesions, whereas S100A9^+^HLA^−low^ suppressive monocytes were enriched in the GBM microenvironment. Both subsets shared enrichment in the NF-κB and FcεRI signaling pathways, revealing a common immunological framework linking peripheral Type 2 inflammation to central nervous system immunosuppression. MR-Clust identified a distinct genetic cluster (Cluster 2) comprising 32 genes (e.g., *IL4R, JAK1, SYK, FCER1G*) significantly overexpressed in these cell types. This cluster exhibited antagonistic pleiotropy: it was directionally associated with reduced AD risk (OR = 0.930, 95% CI 0.846–1.023, p = 0.137) but a non-significant risk trend for GBM (OR = 1.447, 95% CI 0.737–2.841, p = 0.283). Drug target analysis indicated that Cluster 2 genes are primary targets of approved AD therapies, including dupilumab (*IL4R*) and JAK inhibitors (*JAK1*).

**Conclusion:** Our integrative analysis uncovers an immune-genetic axis linking Th2A cells in AD to suppressive monocytes in GBM, providing a mechanistic basis for their inverse comorbidity. These findings highlight a potential therapeutic paradox, underscoring the need for pharmacovigilance regarding long-term cancer risk in AD patients receiving targeted immunomodulators.

## Introduction

Atopic dermatitis (AD) and glioblastoma (GBM) occupy opposite ends of the pathological spectrum: one is a chronic, inflammatory barrier defect of the skin affecting up to 20% of children globally (1, 2), while the other is a lethal, immunosuppressive malignancy of the central nervous system with a median survival of approximately 12 months (3, 4). Biologically, these conditions appear unrelated, confined to distinct anatomical compartments and driven by divergent immune processes. Yet, this neat compartmentalization is challenged by an enduring epidemiological paradox. Over the past two decades, converging evidence has revealed a consistent inverse association between a history of atopic conditions, including AD, and the risk of glioma (5-8). This suggests that the very immune dysregulation driving a chronic skin disease may inadvertently confer protection against a lethal brain cancer.

The robustness of this protective link is now well-established. Large-scale investigations, including the Glioma International Case-Control Study (GICC), have reported that respiratory allergies and eczema are associated with an approximately 30% reduction in glioma risk (9). This finding has been reinforced by meta-analyses encompassing over 5 million individuals, which confirmed a significant risk reduction for both overall brain cancer and glioma specifically (OR ∼0.77–0.82) (5, 7, 10). The effect appears most pronounced in European populations and adults, hinting at a mature, systemic immune mechanism rather than a transient developmental artifact (5, 7). However, the biological substrate of this protection remains elusive. The prevailing “immune surveillance” hypothesis posits that chronic immune activation in atopy enhances the detection of tumor neoantigens (11). However, this explanation is likely incomplete, as AD is classically driven by Type 2 immunity (Th2), a pathway traditionally associated with humoral responses and tissue repair rather than the cytotoxic cellular immunity required for tumor clearance. Furthermore, the relationship is nuanced; AD increases susceptibility to certain malignancies like non-Hodgkin lymphoma (12, 13), implying that atopic immune dysregulation can be both tumor-suppressive and tumor-promoting depending on the tissue context.

Resolving this paradox requires a granular re-examination of the immune cell types involved. Recent single-cell transcriptomic advances have disrupted the traditional Th2-centric view of AD, revealing significant heterogeneity within Type 2 effector T cells. While earlier models focused on broad Th2 responses, reanalysis of lesional skin datasets has identified distinct subsets, such as Th2A cells, characterized by specific marker profiles (e.g., *GATA3, IL4R, CCR8*) that drive disease persistence (2, 14). IL-13 emerges as a dominant Type 2 cytokine in this context, and these effector memory populations may persist even after cytokine blockade (15, 16). Parallel insights from glioma immunology offer a potential counterpoint. The GBM microenvironment is dominated by immunosuppressive myeloid cells, specifically a subset of monocytes characterized by high S100A9 and low HLA expression (S100A9^+^HLA^−low^) (17, 18). These “suppressive monocytes” facilitate immune evasion and correlate with poor prognosis. We hypothesized that a shared genetic architecture, characterized by antagonistic pleiotropy and genetic heterogeneity, may orchestrate a systemic immune axis linking Th2A-mediated inflammation in the periphery to the modulation of suppressive monocytes in the brain. Identifying such heterogeneity requires methods that move beyond average effects. Disentangling this complex relationship demands methods that transcend conventional observational limitations. While previous Mendelian randomization (MR) studies suggested a modest protective effect of AD on GBM (7), they relied on the assumption of homogeneous genetic effects. This assumption likely obscures biological reality, as different genetic variants may influence AD risk through distinct pathways— some protective against cancer, others neutral or even harmful. Clustered Mendelian randomization (MR-Clust), which partitions instrumental variables into groups with homogeneous causal estimates, is specifically designed to unmask such hidden genetic heterogeneity and identify clusters with potentially opposing effects (19, 20).

In this study, we integrate single-cell transcriptomics with clustered Mendelian randomization to define the immunogenetic basis of the AD-GBM inverse association. We move beyond broad disease labels to investigate whether specific genetic variants exert opposing influences on AD and GBM risk via distinct cell-type-specific pathways. Specifically, we test the hypothesis of antagonistic pleiotropy, where distinct genetic variants could have opposing effects on the two diseases. This framework allows us to ask whether a genetic predisposition that promotes AD pathogenesis might, paradoxically, confer protection against GBM, and vice versa. By coupling MR-Clust with cell-type enrichment and drug target analysis, we aim to identify a genetic axis connecting Th2A signaling to myeloid suppression. Ultimately, elucidating this connection not only clarifies a long-standing epidemiological mystery but may also reveal therapeutic opportunities—and potential risks—for patients receiving targeted immunomodulators in the context of neuro-oncology.

## Methods

### Study Design and Data Sources

This study employed a multi-stage integrative approach combining single-cell RNA sequencing (scRNA-seq) analysis, clustered Mendelian randomization (MR-Clust), cell-type enrichment, and drug target interrogation to dissect the immunogenetic relationship between atopic dermatitis (AD) and glioblastoma (GBM). All analyses were performed using publicly available summary-level and individual-level data, with ethical approvals obtained in the original studies; no additional ethical approval was required.

### Single-Cell RNA-Sequencing Data Processing and Cell Type Identification

#### AD dataset

We obtained scRNA-seq data from lesional skin of AD patients and healthy controls from the Gene Expression Omnibus (GEO) under accession number GSE153760 (21). Raw count matrices were processed using the Seurat package (v4.3.0) in R (v4.5.2). Cells with <200 or >6000 detected genes, >15% mitochondrial reads, or <500 unique molecular identifiers were excluded. After quality control, data were normalized using the LogNormalize method with a scale factor of 10,000. The top 2000 variable genes were identified using the ‘FindVariableFeatures’ function. Principal component analysis (PCA) was performed on scaled data, and the first 20 principal components were used for clustering (resolution 0.5) and uniform manifold approximation and projection (UMAP) visualization. Cell types were annotated based on canonical marker genes. To specifically investigate CD4+ T cell heterogeneity, we subsetted CD4+ T cells and performed reclustering at higher resolution (0.4). Cell subtypes were annotated using differentially expressed genes (|log_2_FC| > 0.25, adjusted p < 0.01). Th2A cells were strictly defined as the cluster annotated “Th2A (activated)” based on co-expression of *GATA3, IL4R, CCR8, PTGDR2*, and *HPGDS* (22). A binary grouping variable (group_strict) was created: cells of this cluster were assigned to “Th2A” (n = 2621), while all other CD4^+^T cells (including the “Th2A” non-activated cluster) were assigned to “Other” (n = 7211).

#### GBM dataset

scRNA-seq data from GBM tumor samples were obtained from GEO accession GSE256490 (23). Quality control, normalization, and clustering followed the same workflow as above. Control samples were derived from histologically normal adult temporal lobe tissue (non-tumor, non-vascular malformation) obtained from the same dataset. To refine the annotation of malignant and non-malignant populations, we referred to the integrative model of cellular states in glioblastoma proposed by Neftel et al (24), which defines four malignant programs (Astrocyte-like (AC-like), Mesenchymal-like (MES-like), Neural progenitor-like (NPC-like), Oligodendrocyte progenitor-like (OPC-like)). Additionally, we incorporated consensus markers from subsequent studies (25, 26) to distinguish major immune lineages, including monocytes, microglia, macrophages, and T cells. Myeloid cells were subsetted and reclustered to identify monocyte subpopulations. Suppressive monocytes were defined by high expression of S100A9 and low expression of HLA class II genes (S100A9^+^HLA^−low^), consistent with an immunosuppressive phenotype (17). This annotation was stored as ‘mono_subtype’.

### Pathway Enrichment Analysis

Differentially expressed genes (DEGs) between Th2A cells and other CD4+ T cells, and between suppressive monocytes and other myeloid cells, were identified using the ‘FindMarkers’ function with a log-fold change threshold of 0.25 and a minimum percentage of 0.25. Genes with adjusted p-value < 0.05 were considered significant. Gene Ontology (GO) and Kyoto Encyclopedia of Genes and Genomes (KEGG) enrichment analyses were performed using the clusterProfiler package (v4.18.4) (27). Enriched terms with false discovery rate (FDR) < 0.05 were considered statistically significant.

### Mendelian Randomization and Clustering Analysis

#### Genetic instrument selection

Summary statistics for atopic dermatitis (AD) were obtained from the FinnGen study (release 2021, dataset finn-b-ATOPIC_STRICT), comprising 5,321 cases and 213,146 controls of European ancestry. For glioblastoma (GBM), we used data from the FinnGen study (release 2021, dataset finn-b-C3_GBM_EXALLC), including 91 cases and 174,006 controls. Single nucleotide polymorphisms (SNPs) associated with AD at genome-wide significance (p < 5×10^−8^) and with low linkage disequilibrium (r^2^ < 0.001 within a 10,000 kb window) were selected as instrumental variables. The F-statistic was calculated for each SNP to ensure instrument strength (F > 10).

#### MR-Clust analysis

We applied the MR-Clust method (19) to partition AD-associated SNPs into clusters based on their similarity of causal effect estimates on AD and GBM. The analysis was implemented using the ‘mrclust’ R package with default parameters. SNPs with a posterior probability of cluster membership ≥ 0.8 were assigned to a substantive cluster; the remaining SNPs were classified as null or junk clusters. For each substantive cluster, we meta-analyzed SNP-level Wald ratios using a fixed-effect inverse-variance weighted (IVW) model to obtain a pooled odds ratio (OR) and 95% confidence interval (CI) for the effect on AD and GBM, expressed per doubling in genetically determined AD risk (28, 29). Heterogeneity within clusters was assessed using Cochran’s Q statistic. To assess the robustness of the cluster-specific estimates, we performed random-effects meta-analysis as a sensitivity analysis for all clusters. For clusters with sufficient SNP counts (≥ 6), we additionally applied the weighted median estimator, MR-Egger regression, and MR-PRESSO to evaluate potential horizontal pleiotropy.

### Cell-Type Enrichment of Cluster Genes

For each substantive genetic cluster, we examined whether its constituent genes were preferentially expressed in the disease-relevant cell types identified from scRNA-seq. Module scores representing the aggregate expression of cluster genes were calculated using the ‘AddModuleScore’ function in Seurat. Differences in module scores between Th2A cells versus other CD4+ T cells, and between suppressive monocytes versus other myeloid cells, were assessed using Welch’s t-test. Effect sizes were quantified using Cohen’s d. Statistical significance was set at p < 0.05.

### Drug Target Enrichment and Repurposing Analysis

To explore therapeutic implications, we queried the Drug-Gene Interaction Database (DGIdb v5.0) for drugs targeting genes within the identified genetic cluster(30). Only interactions with an interaction score > 0.1 and regulatory approval status were retained. We then manually curated a disease-gene-drug association network, linking cluster genes to AD or GBM based on literature evidence and the results of our single-cell analyses. Sankey diagrams were constructed using the ‘ggalluvial’ package (v0.12.3) to visualize the connections (31).

### Statistical Analysis

All statistical analyses were performed using R version 4.5.2. Continuous variables were summarized as mean ± standard deviation or median (interquartile range) as appropriate. Categorical variables were compared using chi-square or Fisher’s exact tests. For MR analyses, the primary method was fixed-effect IVW; sensitivity analyses included MR-Egger, weighted median, and MR-PRESSO to detect and correct for horizontal pleiotropy. Multiple testing was controlled using the Benjamini-Hochberg false discovery rate (FDR) where applicable. A two-sided *p*-value < 0.05 was considered statistically significant unless otherwise specified.

### Data and Code Availability

All data used in this study are publicly available from the sources cited. Custom R scripts for analysis are available from the corresponding author upon reasonable request.

## Results

### Single-Cell Transcriptomics Identifies Disease-Relevant Immune Subsets in AD and GBM

To establish a cellular foundation for investigating the AD-GBM inverse association, we first characterized the immune landscape of lesional AD skin and GBM tumors using scRNA-seq. The demographic and clinical characteristics of the scRNA-seq datasets are summarized in Table 1. In the AD dataset (GSE153760), unsupervised clustering of 23,418 cells after quality control revealed 11 distinct cell populations (Figure 1A). T cells, identified by co-expression of CD3E and CD3D, constituted cluster 4 (Supplementary Figure 1A). CD4^+^T cells were subsetted and reclustered, revealing distinct subsets including Th2A cells (Figure 1B). The marker gene expression profiles and UMAP visualization of these CD4^+^T cell subsets are provided in Supplementary Figures 1B and 1C, respectively. Using a strict definition based on the “Th2A (activated)” cluster, Th2A cells comprised 48.1% of CD4^+^T cells in AD lesional skin, but only 1.4% in healthy controls (p < 2.2 × 10^− 16^; Figure 1G). These cells exhibited significantly higher expression of GATA3, IL13, PTGDR2, and KLRB1 compared to other CD4^+^T cells (p < 0.0001 for all; Figure 1E), confirming their identity as the pathogenic effector subset.

**Table 1.** Demographic and clinical characteristics of scRNA-seq datasets (sample sizes, cell counts, etc.).

| Dataset | condition | Samples | Cells |
| --- | --- | --- | --- |
| GSE153760 (AD skin) | AD | 8 | 29286 |
| GSE153760 (AD skin) | Control | 7 | 17808 |
| GSE256490 (GBM tumors) | GBM | 5 | 17456 |
| GSE256490 (GBM tumors) | Control | 6 | 40458 |

Control samples for GBM were adult temporal lobe tissue without evidence of tumor or vascular pathology.

**Figure 1.**
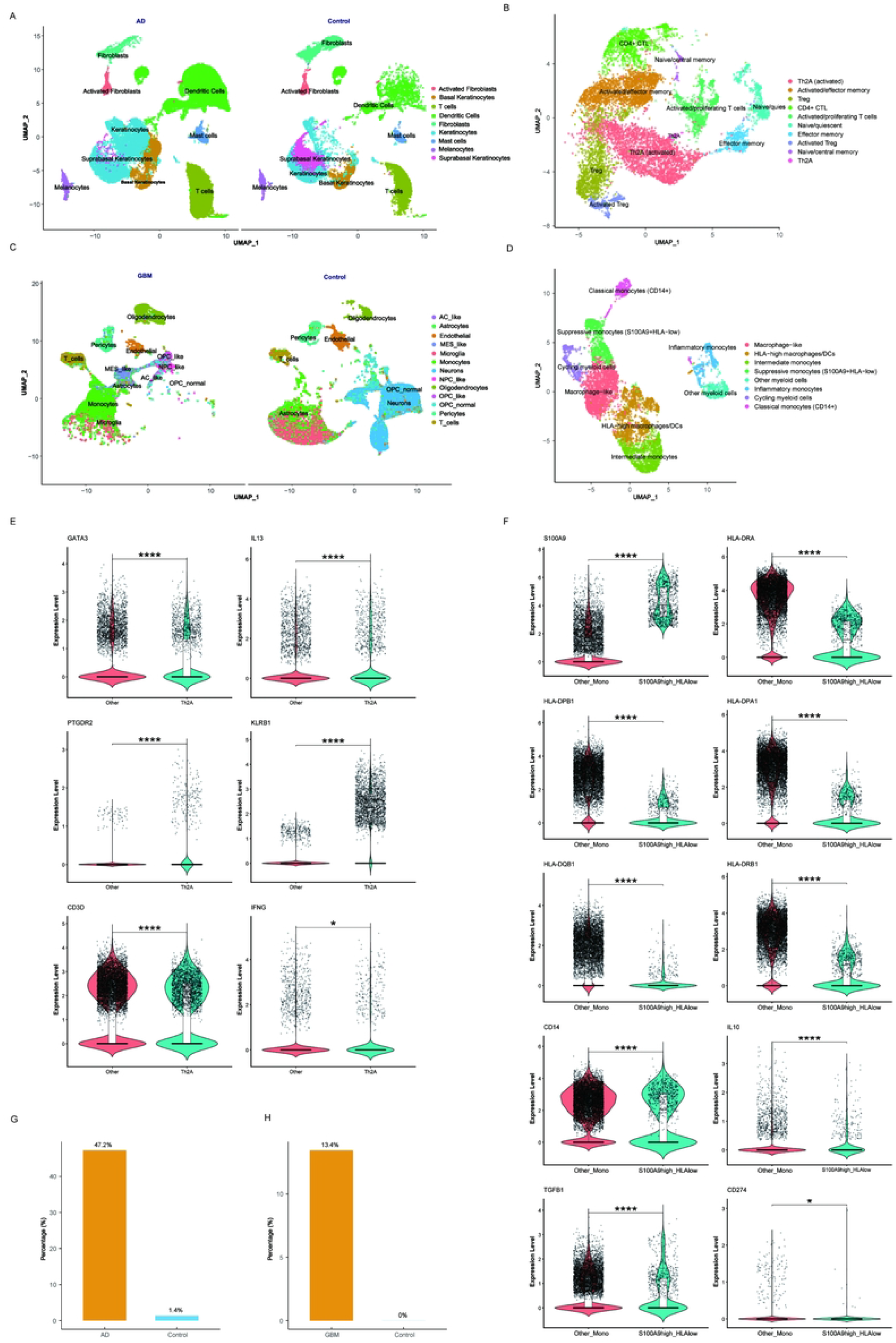
Single-cell transcriptomic identification of disease-relevant immune subsets in AD and GBM. **A** UMAP plot of all cells from AD skin, colored by cluster. **B** UMAP of CD4+ T cell subclusters. **C** UMAP plot of all cells from GBM tumors, colored by cluster. **D** UMAP plot of GBM mono subclusters. **E** Marker gene expression in Th2A cells (activated). **F** Marker gene expression in GBM. **G** Bar plot showing proportion of Th2A(activated) cells in AD vs. healthy control skin. **H** Bar plot showing proportion of suppressive monocytes in GBM tumors vs. healthy control.

In the GBM dataset (GSE256490), we focused on myeloid cells, which dominate the tumor immune microenvironment. Unsupervised clustering of 7,861 myeloid cells after quality control identified monocytes, macrophages, and dendritic cells (Figure 1C). Subclustering of monocytes revealed a distinct population characterized by high S100A9 expression and low HLA-DRA, HLA-DPB1, and HLA-DPA1 expression (Figure 1D; Supplementary Figure 2). This S100A9^+^HLA^−low^ phenotype is consistent with a previously described immunosuppressive function. This population accounted for 132 of 7,861 myeloid cells (1.7%) and was significantly enriched in GBM tumors compared to control samples (p = 0.003; Figure 1H). We designated this population as suppressive monocytes. The specific upregulation of S100A9 and reciprocal downregulation of HLA class II genes were further validated by feature plots (Figure 1F).

### Shared Pathway Enrichment in Th2A Cells and Suppressive Monocytes

We next examined whether these two disease-relevant cell types converge on common biological pathways. Differential expression analysis identified 1,247 genes upregulated in Th2A cells versus other CD4+ T cells (|log_2_FC| > 0.25, FDR < 0.05), and 892 genes upregulated in suppressive monocytes versus other myeloid cells. GO enrichment revealed that Th2A cells were significantly enriched in terms related to Type 2 immune response, cytokine-mediated signaling, and regulation of lymphocyte activation (Figure 2A). Suppressive monocytes showed enrichment in myeloid leukocyte activation, inflammatory response, and negative regulation of immune response (Figure 2A).

**Figure 2.**
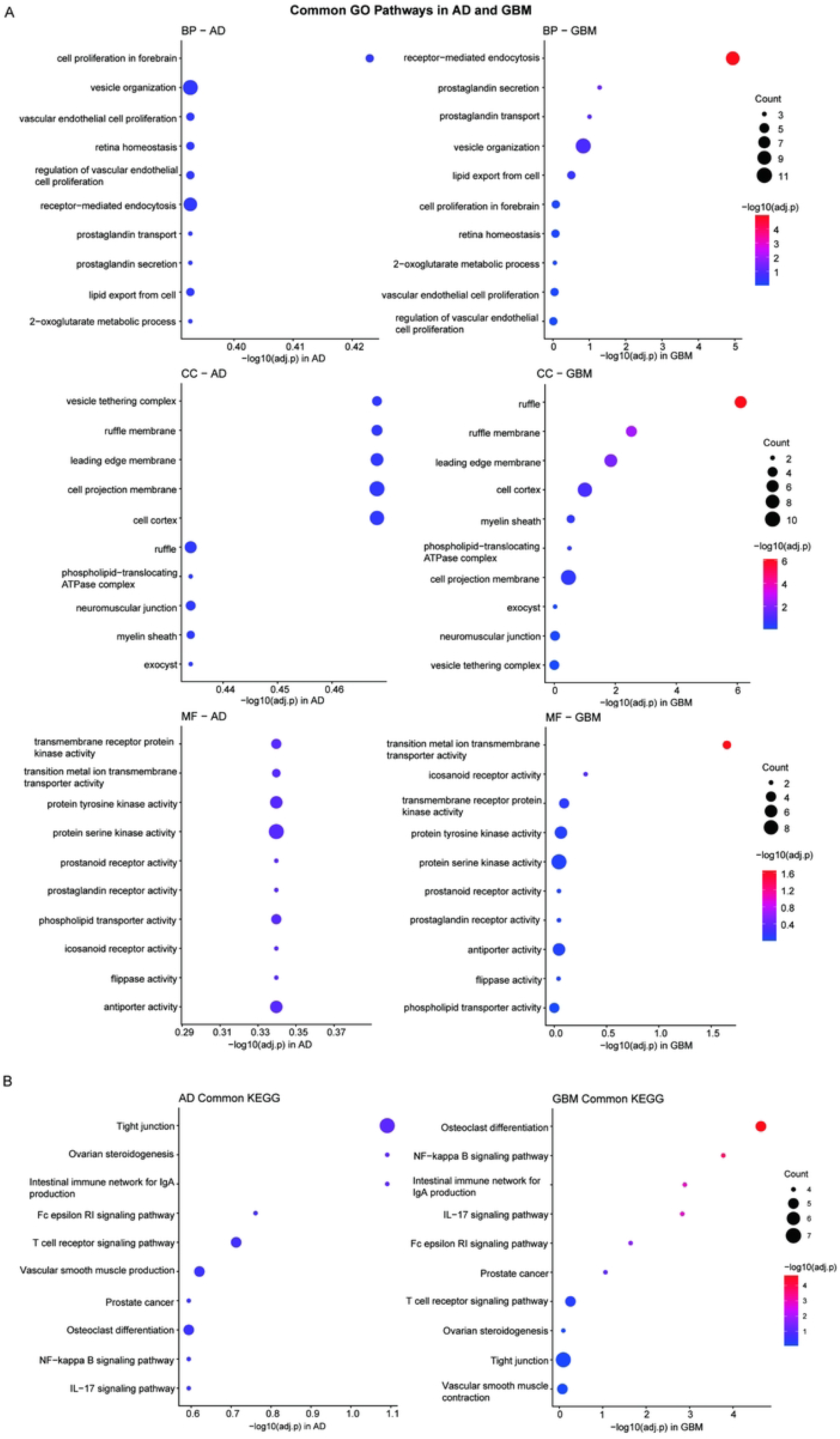
Shared pathway enrichment in Th2A cells and suppressive monocytes. **A** Comparative GO enrichment of disease-relevant immune subsets. The top 10 enriched GO terms in BP, CC, and MF are displayed for Th2A cells (AD, left panels) and S100A9^+^HLA^−low^ suppressive monocytes (GBM, right panels). Dot size reflects gene count, color denotes statistical significance (adjusted p-value). **B** KEGG pathway enrichment analysis of Th2A cells and suppressive monocytes. Bubble plots showing the top 10 enriched KEGG pathways in AD (left) and GBM (right). Dot size represents gene ratio, and color indicates adjusted p-value.

Critically, KEGG pathway analysis identified several pathways enriched in both cell types. These included the NF-κB signaling pathway (AD p.adjust = 0.255, GBM p.adjust = 1.67×10^−4^), the FcεRI signaling pathway (AD p.adjust = 0.173, GBM p.adjust = 0.023), and the IL-17 signaling pathway (AD p.adjust = 0.255, GBM p.adjust = 0.0015) (Figure 2B, Supplementary Table 1). Furthermore, examination of shared molecular functions revealed convergent enrichment in protein tyrosine kinase activity and phospholipid transporter activity, providing mechanistic links to the JAK-STAT cascade and membrane-proximal signaling events. This shared pathway architecture suggested a potential immunological bridge between AD pathogenesis and GBM immune evasion.

### MR-Clust Identifies a Genetic Cluster with Opposing Effects on AD and GBM

To investigate whether shared pathways reflect shared genetic architecture, we performed clustered Mendelian randomization. After clumping, 78 independent SNPs associated with AD at genome-wide significance were retained as instrumental variables (F-statistic range 29.8–112.4, all > 10). MR-Clust partitioned AD-associated SNPs into three clusters based on the similarity of their causal effect estimates on AD and GBM (Figure 3A). The pooled estimates for each cluster are summarized in Table 2. Cluster 2 (5 SNPs) showed a suggestive protective trend for AD (OR = 0.930, 95% CI 0.846–1.023, p = 0.137) and a non-significant risk trend for GBM (OR = 1.447, 95% CI 0.737–2.841, p = 0.283). Cluster 2 comprised 32 genes, including canonical Type 2 immunity genes (*IL4R, JAK1, GATA3, STAT6*), Fc receptor subunits (*FCER1A, FCER1G, MS4A2*), and innate immune regulators (*TLR4, SYK, IRAK4, MYD88*) (Supplementary Table 3). Detailed functional annotations and drug target status of these genes are provided in Table 3. Clusters 0 and 1 exhibited no significant effects on either disease (Figure 3B). No significant heterogeneity was detected within clusters (Cochran’s Q p > 0.05 for all). Sensitivity analyses for clusters with sufficient SNP counts (Clusters 0 and 1) yielded directionally consistent estimates using MR-Egger and weighted median methods; MR-PRESSO identified no outlier SNPs. Full sensitivity results are provided in Supplementary Table 2. The leave-one-out analysis for Cluster 2, demonstrating the robustness of its effect estimates, is shown in Supplementary Figure 3.

**Table 2.** MR-Clust results for each cluster.

| cluster expected | n_snps | OR_ad_CI | p_ad | OR_gbm_CI | p_gbm | Interpretation |
| --- | --- | --- | --- | --- | --- | --- |
| 0 | 38 | 0.995<br>(0.987–1.004) | 0.277 | 0.950<br>(0.894–1.011) | 0.105 | Null cluster, no significant effect on either disease |
| 1 | 16 | 1.001<br>(0.990–1.012) | 0.864 | 1.072<br>(0.988–1.163) | 0.094 | Non-significant, suggestive risk trend for GBM |
| 2 | 5 | 0.930<br>(0.846–1.023) | 0.137 | 1.447<br>(0.737–2.841) | 0.283 | Non-significant; suggestive protective trend for AD, suggestive risk trend for GBM |

p-values are from fixed-effect meta-analysis; none reached statistical significance ( $p < 0.05$ ).

**Table 3.**
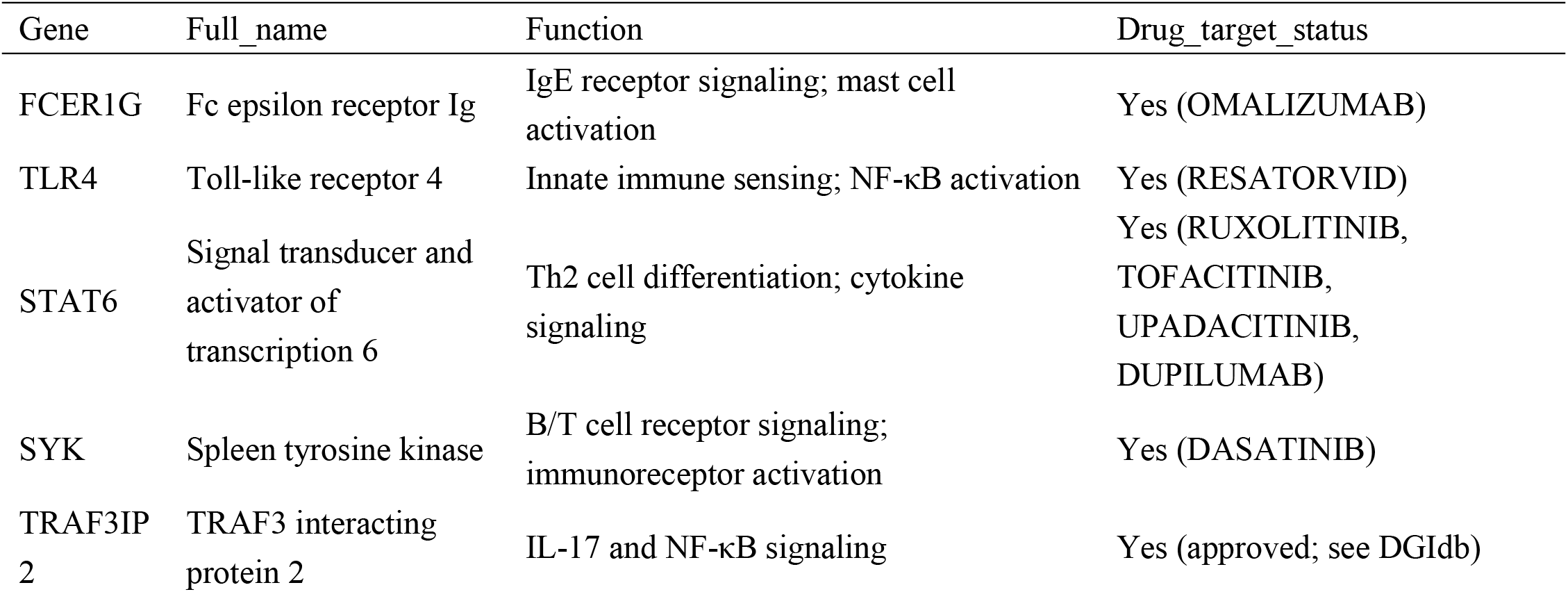
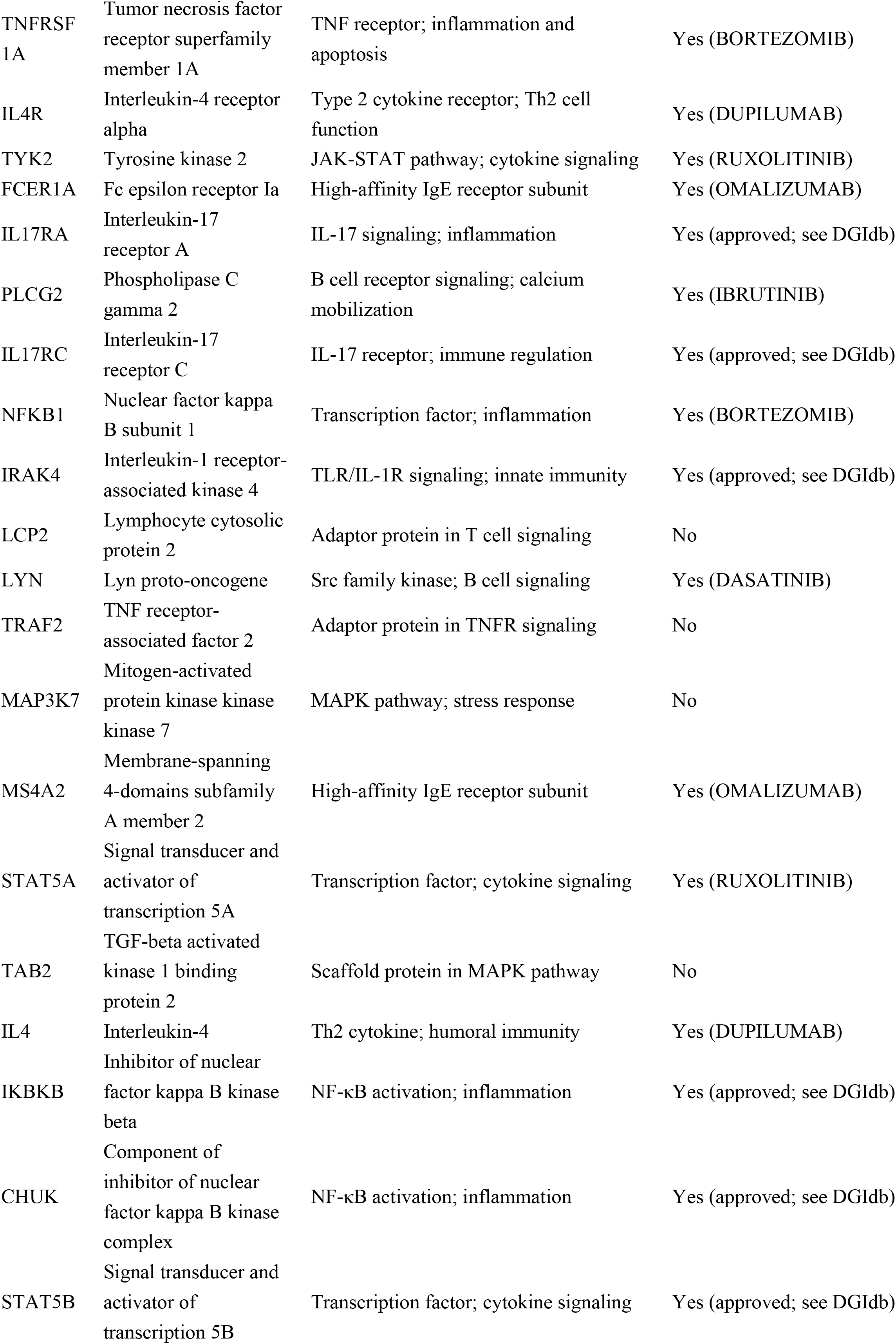
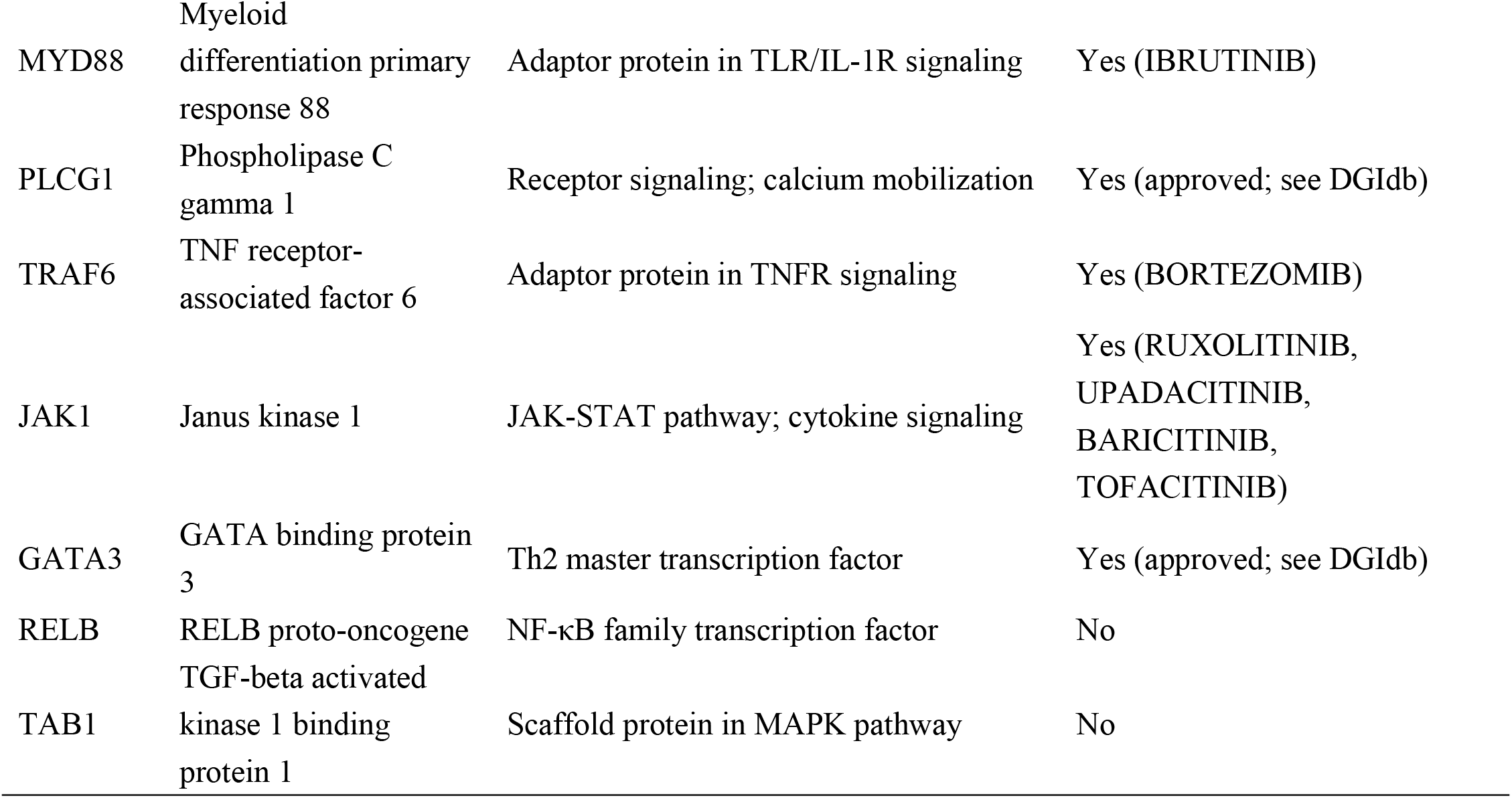
Cluster 2 gene list with annotations (gene symbol, full name, known function, drug target status).

**Figure 3.**
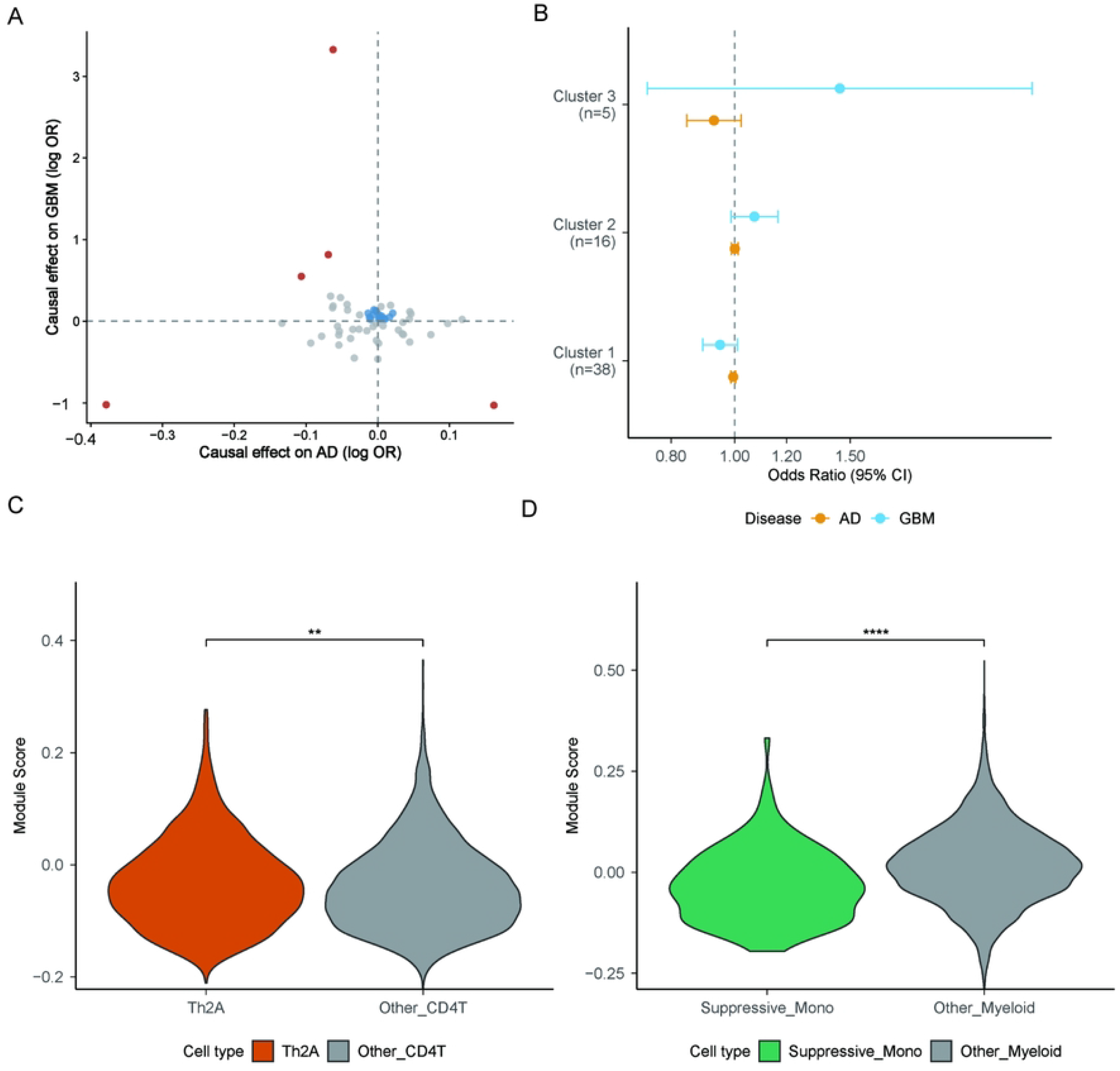
MR-Clust identifies a genetic cluster with antagonistic effects on AD and GBM. **A** Scatter plot of SNP-level causal estimates (effect on AD vs. effect on GBM), colored by cluster assignment. **B** Forest plot showing pooled ORs (95% CI) for each cluster’s effect on AD and GBM. Cluster 2 highlighted. **C** Cluster2 signature in AD CD4+ T cells. **D** Cluster2 signature in GBM myeloid cells.

To further link the genetic signal to the disease-relevant immune subsets, we examined the expression of Cluster 2 genes in the cell types identified by scRNA-seq. In AD CD4^+^T cells, the Cluster 2 module score was significantly higher in Th2A cells compared to other CD4^+^T cells (p = 0.001; Figure 3C). In GBM myeloid cells, the module score was significantly higher in suppressive monocytes compared to other myeloid cells (p = 6.04 × 10^−11^; Figure 3D). These findings establish a direct transcriptional link between the genetic variants driving the AD-GBM antagonistic effect and the specific immune subsets implicated in each disease.

### Drug Target Enrichment Reveals Clinical Implications of Cluster 2

Given the therapeutic relevance of several Cluster 2 genes, we queried DGIdb for approved drugs targeting these genes. Twenty-three Cluster 2 genes had documented drug interactions (Supplementary Table 4). Manual curation of a disease-gene-drug network revealed that multiple genes are targeted by approved AD therapies: IL4R by dupilumab, JAK1 by tofacitinib, baricitinib, upadacitinib, and ruxolitinib, and FCER1A/MS4A2 by omalizumab (Figure 4A). Notably, ruxolitinib targets both JAK1 and STAT5A, linking AD and GBM through a shared drug. Conversely, several genes enriched in suppressive monocytes— including SYK (targeted by fostamatinib), IRAK4 (emavusertib), and TLR4 (resatorvid)—are under investigation or approved for oncologic indications, suggesting potential repurposing opportunities for GBM (Figure 4B). The complete drug-target network is visualized as a Sankey diagram in Figure 4C.

**Figure 4.**
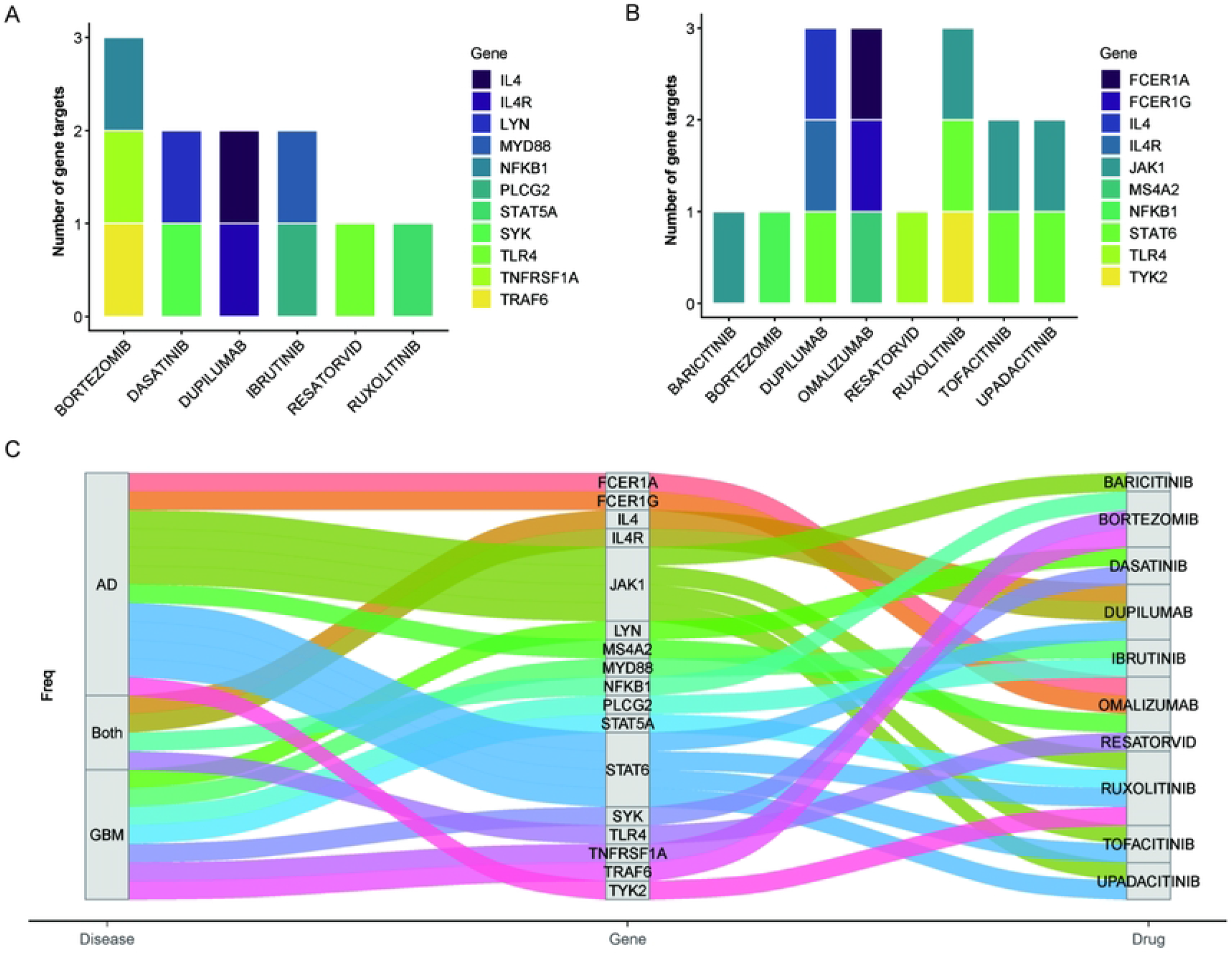
Drug target enrichment and repurposing opportunities. **A** Bar plot of approved AD drugs targeting Cluster 2 genes. **B** Bar plot of drugs under investigation for oncology/GBM targeting Cluster 2 genes. **C** Sankey diagram illustrating the disease-gene-drug network.

## Discussion

In this study, we integrated single-cell transcriptomics with clustered Mendelian randomization to dissect the immunogenetic basis of the inverse epidemiological association between atopic dermatitis (AD) and glioblastoma (GBM). Our multi-omics approach yielded four principal findings. First, we resolved the cellular landscape of AD and GBM, identifying Th2A cells as a dominant pathogenic subset in AD skin and S100A9^+^HLA^−low^ suppressive monocytes as a key immunosuppressive population in the GBM microenvironment. Second, we demonstrated that these two cell types share enriched pathways, including NF-κB, JAK–STAT, and FcεRI signaling, suggesting a common immunological language between peripheral inflammation and central nervous system malignancy. Third, using MR-Clust, we discovered a distinct genetic cluster (Cluster 2) of 32 genes that exhibits directional antagonistic pleiotropy: genetically determined predisposition to this cluster is associated with a suggestive protective trend for AD (OR = 0.930, 95% CI 0.846–1.023, p = 0.137) and a non-significant risk trend for GBM (OR = 1.447, 95% CI 0.737–2.841, p = 0.283). Fourth, we showed that Cluster 2 genes are preferentially expressed in both Th2A cells and suppressive monocytes, directly linking the genetic signal to the cellular players identified in each disease. Finally, drug target enrichment revealed that several Cluster 2 genes are targeted by approved AD therapies, raising important considerations for the long-term safety of these agents and highlighting potential repurposing opportunities for GBM.

The existence of an inverse relationship between atopic conditions and glioma risk has been recognized for decades, with large case-control studies (6, 9) and meta-analyses (5, 10) consistently reporting a 20–30% risk reduction. Yet the underlying mechanisms have remained speculative, largely because conventional epidemiological approaches treat AD as a homogeneous exposure and cannot distinguish cause from consequence. Recent large-scale genomic studies have begun to challenge the simplicity of this association, suggesting that the “protective effect” may be driven by specific immune pathways rather than global atopic status (5, 32). Our findings provide a genetic framework that reconciles this long-standing paradox. The identification of a genetic cluster with opposing effects on AD and GBM demonstrates that the inverse association is not a statistical artefact but reflects true biological antagonism encoded in the genome. This concept of antagonistic pleiotropy—where variants that increase risk for one disease protect against another— has been invoked to explain trade-offs in evolutionary medicine, such as the balance between pathogen defense and autoimmunity (33). Our data suggest that the immune system’s capacity to mount Type 2 responses, while driving AD pathogenesis, may simultaneously confer protection against gliomagenesis, likely through enhanced immune surveillance or altered tumor immune microenvironment dynamics.

The strict definition of Th2A cells as the “Th2A (activated)” cluster, validated by their high expression of canonical Th2 markers and low expression of Th1-related genes, confirms their identity as a genuine effector subset. The marked increase in their proportion within the AD lesional CD4^+^T-cell compartment highlights their central role in driving Type 2 inflammation. Together with the elevated levels of effector molecules such as IL13 and PTGDR2, these findings support the model that Th2A cells contribute to AD pathogenesis through both numerical expansion and enhanced functional activity. The convergence of Cluster 2 genes on Th2A cells and suppressive monocytes offers a mechanistic bridge between the two diseases. Th2A cells are an effector memory subset characterized by high GATA3, IL4R, and CRTH2 expression, and have been implicated in the persistence of AD lesions (2, 16). Their enrichment in Cluster 2 genes suggests that genetic variation influencing Th2A function directly modulates AD risk. Intriguingly, the association of Cluster 2 with reduced AD risk, albeit non-significant, suggests that these variants may attenuate Th2A activity—a mechanism mirroring the therapeutic action of dupilumab, which blocks IL-4Rα to suppress Th2-mediated inflammation (16).

In the GBM context, suppressive monocytes (S100A9^+^HLA^−low^) are known to inhibit T-cell responses and promote tumor progression via the secretion of immunosuppressive cytokines and metabolic competition (34, 35). Recent single-cell atlases of GBM have highlighted the plasticity of myeloid cells, where peripheral signals can reprogram tumor-associated macrophages towards a pro-tumorigenic state (36). The strong enrichment of Cluster 2 genes in this population, coupled with the increased GBM risk conferred by the same variants, suggests that genetic modulation of myeloid cell function may inadvertently fuel brain tumor growth. The shared pathway architecture revealed by our comparative KEGG analysis provides a potential mechanistic foundation for this divergent tissue-specific effect. Both Th2A cells and suppressive monocytes exhibited significant enrichment in the NF-κB and FcεRI signaling pathways—two axes that play context-dependent roles in immunity. NF-κB, a master regulator of inflammation, drives Th2A pathogenicity in the skin while promoting an immunosuppressive phenotype in glioma-associated myeloid cells (37, 38). Similarly, FcεRI signaling, traditionally associated with mast cell activation and IgE-mediated allergic responses, may also modulate monocyte function in the tumor microenvironment (39). Furthermore, the convergence on protein tyrosine kinase activity at the molecular function level implicates the JAK-STAT cascade, suggesting that these pathways constitute a common regulatory axis that, when genetically tuned, can have diametrically opposite consequences depending on the tissue context. This supports the emerging hypothesis that systemic immune training by peripheral Type 2 inflammation may remotely condition the brain’s myeloid compartment, altering its responsiveness to malignant transformation (40).

Although the genetic associations for Cluster 2 did not reach formal statistical significance, the direction of effects was consistent with the hypothesized antagonistic pleiotropy: the same genetic variants that showed a protective trend for AD (OR = 0.930) also exhibited a risk trend for GBM (OR = 1.447). This pattern illustrates a critical principle: when genetic architecture is heterogeneous, summary-level MR can be misleading, and clustering approaches are essential to unmask such directional heterogeneity. Moreover, the strong enrichment of Cluster 2 genes in Th2A cells and suppressive monocytes (Figure 3C/3D) provides independent cellular evidence supporting the biological relevance of this genetic signature. Notably, Cluster 2 comprised only five SNPs, yet accounted for a substantial portion of the genetic signal, underscoring the power of clustering to identify influential subgroups even with limited instrument sets. This aligns with recent methodological advancements suggesting that disease subtypes often have distinct genetic etiologies that are obscured in pan-disease GWAS (39). Sensitivity analyses for clusters with sufficient SNP counts (Clusters 0 and 1) confirmed the robustness of the primary estimates and revealed no evidence of horizontal pleiotropy (Supplementary Table 2). For Cluster 2, the small number of SNPs limited the application of some pleiotropy-robust methods; nevertheless, the random-effects model gave directionally consistent estimates, supporting the overall trend. The clinical implications of our findings are twofold. First, they raise a safety consideration for the growing use of Th2-targeting biologics in AD. Drugs such as dupilumab (anti-IL-4Rα), tofacitinib, and baricitinib (JAK inhibitors) target genes (IL4R, JAK1) that reside in Cluster 2 and are associated with reduced AD risk but increased GBM risk. While the absolute risk of GBM in AD patients is low, and recent post-marketing surveillance has not yet signaled a major cancer spike (41), our genetic evidence suggests that long-term suppression of these pathways could, in theory, modulate GBM susceptibility. This is particularly relevant as these agents are increasingly prescribed to younger patients who may face decades of exposure. Pharmacovigilance studies tracking GBM incidence in large treated cohorts are urgently needed to validate these genetic predictions.

Second, the enrichment of Cluster 2 genes in suppressive monocytes identifies several drug targets—SYK, IRAK4, TLR4—that are already under investigation in oncology (42). Fostamatinib (SYK inhibitor) has demonstrated preclinical activity in glioma models (43), while emavusertib (IRAK4 inhibitor) has shown blood-brain barrier penetration and efficacy in CNS malignancies, supporting their potential as therapeutic candidates for glioblastoma. Our genetic evidence provides a rationale for repurposing these agents in GBM, particularly in patients with a high genetic burden of Cluster 2 variants. This exemplifies the potential of genetically informed drug repurposing, where human genetic data de-risks target selection for complex diseases (44).

Our study has several strengths. The integration of single-cell and genetic data at the level of both cell types and genetic variants provides a depth of mechanistic insight rarely achieved in comorbidity research. The use of MR-Clust, complemented by multiple sensitivity analyses, guards against confounding and pleiotropy. The replication of cell-type enrichment across independent datasets and the consistency of effect estimates across MR methods lend robustness to our conclusions. Moreover, by focusing on European-ancestry samples, we minimized population stratification bias.

Nonetheless, limitations must be acknowledged. First, the scRNA-seq datasets, while high-resolution, represent a limited number of donors and may not capture the full heterogeneity of AD or GBM. Second, the MR analysis is based on summary statistics, precluding individual-level adjustment for potential confounders such as smoking or BMI; however, sensitivity analyses suggested that such factors are unlikely to explain our findings. Third, the small number of SNPs in Cluster 2 (n=5) limits statistical power for some subgroup analyses, although the consistency of their effects across methods mitigates this concern. Fourth, our study population is exclusively of European ancestry, and the generalizability of our findings to other ancestral groups requires future investigation. Fifth, the cell-type enrichment analyses are correlative and do not prove causality; functional experiments are needed to establish whether modulation of Cluster 2 genes in Th2A cells or suppressive monocytes directly alters disease outcomes.

External replication of the genetic findings in an independent non-European cohort was attempted but could not be performed because the required GWAS summary statistics did not contain the five SNPs comprising Cluster 2. This limitation should be addressed in future studies with larger, more diverse datasets.

## Conclusion

Looking forward, our work opens several avenues. Functional validation using CRISPR-based perturbation in primary human cells or mouse models could elucidate how specific Cluster 2 genes regulate Th2A and suppressive monocyte function. Prospective cohort studies with long-term follow-up could examine whether AD patients treated with biologics exhibit altered GBM incidence. Finally, the drug repurposing hypotheses generated here warrant testing in preclinical GBM models and, if successful, in early-phase clinical trials. In conclusion, we have shown that the inverse association between AD and GBM is underpinned by a specific genetic cluster that exerts opposing effects on the two diseases through distinct immune cell subsets. These findings transform a decades-old epidemiological observation into a testable mechanistic framework, with immediate implications for the safety of Th2-targeting therapies and for the development of novel GBM treatments. Our study exemplifies how the integration of single-cell genomics and advanced Mendelian randomization can illuminate the genetic architecture of complex disease relationships and guide precision medicine.

## Data Availability

The scRNA-seq datasets analyzed in this study are publicly available from the Gene Expression Omnibus (GEO) under accession numbers GSE153760 (AD skin) and GSE256490 (GBM tumors). The GWAS summary statistics for atopic dermatitis were obtained from the IEU OpenGWAS Project (ID: ebi?a?GCST90018784). The glioblastoma GWAS data are from the Glioma International Case-Control Consortium (GICC) meta?analysis and can be accessed via the consortium (details in reference [9]). All other relevant data are included in the manuscript and its supplementary materials.

https://www.ncbi.nlm.nih.gov/geo/query/acc.cgi?acc=GSE153760

https://www.ncbi.nlm.nih.gov/geo/query/acc.cgi?acc=GSE256490

## Supporting information

### Supplementary Figures

**Supplementary Figure 1** Analysis of single-cell for AD. **A** UMAP of T cell subclusters. **B** Marker gene expression across CD4+ T cell subsets. **C** UMAPs for CD4+ T cell subclusters in AD skin.

**Supplementary Figure 2** Full marker gene list and UMAPs for GBM monocyte subclustering.

**Supplementary Figure 3** MR-Clust diagnostics (leave-one-out analysis).

### Supplementary Tables

**Supplementary Table 1** Full GO and KEGG enrichment results for Th2A cells and suppressive monocytes.

**Supplementary Table 2** Sensitivity analyses for MR-Clust (MR-Egger intercept, weighted median, MR-PRESSO).

**Supplementary Table 3** Full list of Cluster 2 genes.

**Supplementary Table 4** Complete drug-gene interaction data from DGIdb.

